# Systematic analysis of tumor-infiltrating immune cells in human endometrial cancer: a retrospective study

**DOI:** 10.1101/19003707

**Authors:** Xi Zhou, Zhengjiang Ling, Bing Yang

## Abstract

**Objective:** The prognostic effect of tumor-infiltrating immune cells (TIICs) on endometrial cancer (EMC) has not been extensively investigated. In the present study, we systematically analyzed the role of TIICs in EMC development.

**Methods:** Patient data were downloaded from The Cancer Genome Atlas (TCGA). We comprehensively analyzed TIIC population in EMC tissue and their role in EMC progression and prognosis by using a deconvolution algorithm (CIBERSORT) and clinically annotated expression profiles.

**Results:** The proportions of gamma delta T cells, resting NK cells, M1 macrophages, and resting mast cells were significantly different in normal endometrium and EMC tissue. The proportion of CD8+ T cells, resting memory CD4 T cells, and M0 macrophages was reversed middle correlated. The proportion of resting dendritic cells, resting memory CD4 T cells, and T regulatory cells (Tregs) decreased in accordance with the cancer cell differentiation grade (G); the lower proportion of activated dendritic cells and gamma delta T cells and higher proportion of Tregs predicted longer EMC survival time and vice versa. The low proportion of gamma delta T cells indicated better response to therapy.

**Conclusion:** Collectively, our data suggested subtle differences in the cellular composition of TIICs in EMC, and these differences were likely to be important determinants of both prognosis and therapy of EMC.

## Introduction

Endometrial cancer (EMC) is the most common gynecological malignancy, and approximately 12,160 deaths due to uterine cancer were estimated to occur in the United States in 2019^1^. EMC is the second most common female malignancy in China next to cervical cancer^2^. An advanced stage (III or IV) of EMC at the time of diagnosis is lethal, the percentage of five-years survival time is 43–67% for stage III disease and only 13–25% for stage IV disease^1^. In addition to standard care, targeted therapies specific to individualized tumors, such as immunotherapy, are needed for patients in advanced stages^3^. This prompted researchers to assess the use of tumor-infiltrating immune cells (TIICs) for immunotherapy. Svetlana *et al* demonstrated that infiltration of CD8+ T cells in the tumor epithelium at the invasive border is a favorable prognostic factor for patients with EMC^4^. Pater *et al* reported a decrease in intraepithelial CD3+ tumor-infiltrating lymphocyte (TIL) counts was associated with advanced stage and high risk in patients with EMC^5^. A high proportion of CD8+ PD-1+ lymphocytes was associated with improved prognosis in patients with high-risk EMC^6^. Yamagami *et al* found a high level of regulatory T cells (Tregs) was associated with poorer disease-free survival^7^. These findings suggest that molecular modifiers of the local tumor immune response may be tumor type-specific^8^. Because there is a lack of comprehensive analysis of immune cells in EMC, we systematically analyzed the expression of 22 types of immune cells in EMC and its association with tumor cell grade, clinical stage, and their effects on patient survival time by using data from The Cancer Genome Atlas (TCGA).

## MATERIALS AND METHODS

### Data acquisition

The data for this study were downloaded from TCGA on July 10, 2019. Patients with any missing or insufficient data regarding tumor cell grade, clinical stages (FIGO stage), survival time, and disease-free survival time were excluded from the subsequent analysis. Preprocessing and aggregation of raw data were normalized using the “limma” package of R software. Details of the study design and the samples included at each stage of analysis are illustrated as a flowchart in Figure 1.

**Figure 1.**
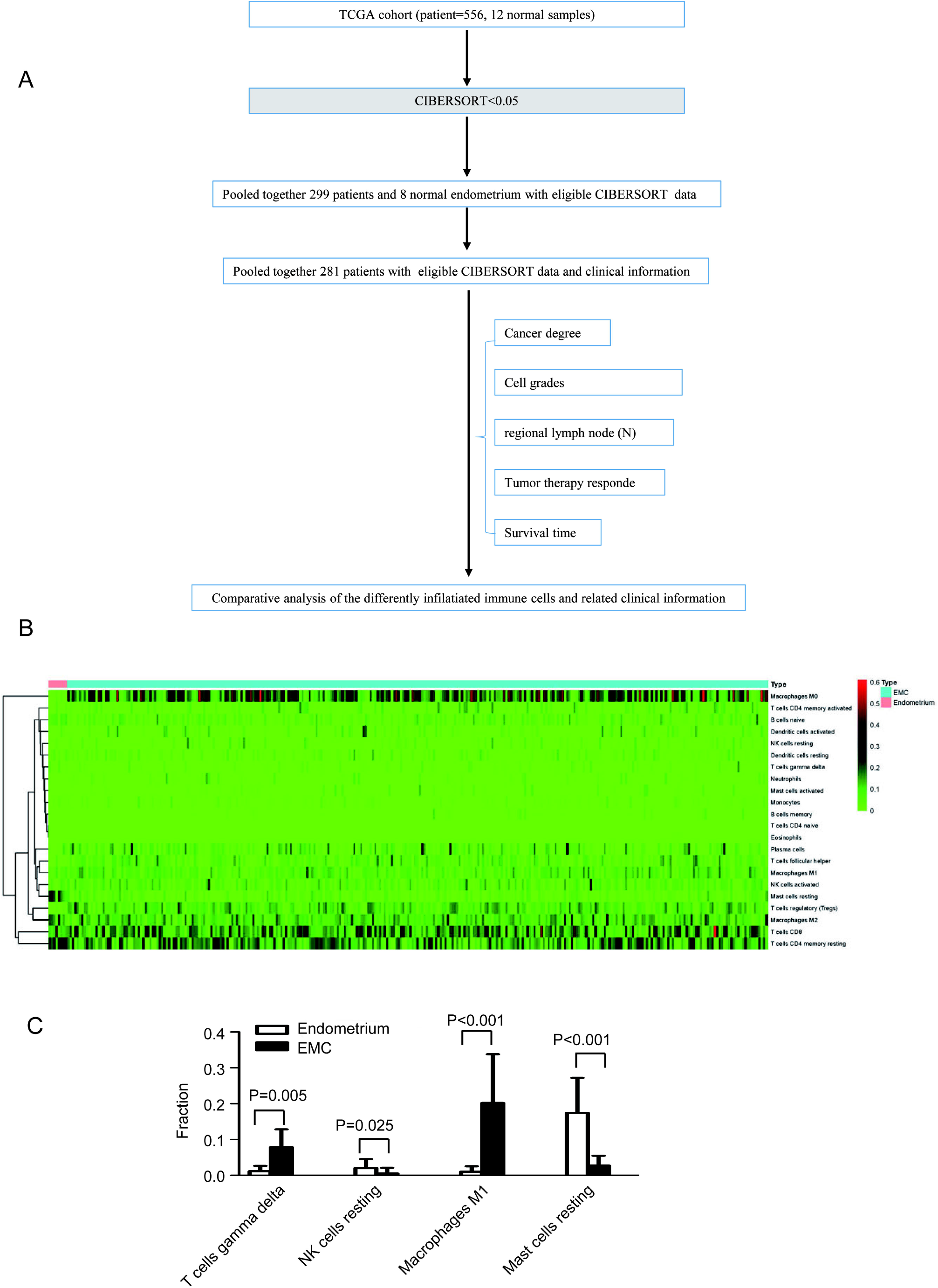
Study flow chart and the proportion of TIICs in normal endometrium and endometrial cancer (EMC) tissue. A, Study flowchart detailing the flow of samples at each stage of analysis. TGCA, The Cancer Genome Atlas. B, The proportion of TIICs in normal endometrium and EMC tissue as determined by a heat map. C, The difference in TIICs between normal endometrium and EMC tissue.

### Evaluation of TIICs

The normalized gene expression data were used to infer the relative proportions of 22 types of TIICs by using the CIBERSORT algorithm as previously reported^9-12^. Briefly, gene expression datasets were prepared using standard annotation files, and the data uploaded to the CIBERSORT web portal (http://cibersort.stanford.edu/), with the algorithm run using the default signature matrix at 1000 permutations^10^. CIBERSORT calculates a P-value for the deconvolution of each sample by using Monte Carlo sampling, thus providing a measure of confidence in the results.

### Statistical analyses

Cases with a CIBERSORT P value of <0.05 were included in the main survival analysis. Associations regarding tumor cell differentiation grade, clinical stage, patient survival time, and the estimated proportions of immune cell types were tested using the Cox regression analysis. Immune cell subsets that were significantly associated with outcome in unadjusted analyses were included in the multivariate models. Multivariate analyses were adjusted for survival time, tumor cell differentiation grades, and clinical stage. The association between infiltrated immune cells and the corresponding disease-free survival time was analyzed by Kaplan-Meier survival curves and evaluated using the log-rank test.

All analyses were conducted using R version 3.5. All statistical tests performed were two-sided, and P values of <0.05 were considered to be statistically significant.

## Results

### Performance of CIBERSORT for characterizing TIIC composition in normal endometrium and EMC tissue

The CIBERSORT analysis result showed that one lymphocyte population (CD4 naïve T cells) was not observed in EMC tissue and four lymphocyte populations (memory B cells, CD4 naïve T cells, activated mast cells, and eosinophils) (Figure 1B) were observed in EMC tissue. The proportions of gamma delta T cells, resting NK cells, M1 macrophages, and resting mast cells were significantly different in normal endometrium and EMC tissue (Figure 1C).

### The correlation of TIICs in EMC

We analyzed the correction of TIICs in CCs. Cytolytic activity was middle reversed correlated with the proportion of CD8+ T cells and resting memory CD4 T cells (Pearson’s correlation = −0.51), CD8+ T cells, and M0 macrophages (Pearson’s correlation = −0.59) in the TCGA cohort at a CIBERSORT P value of <0.05 (Figure 2). Several studies have confirmed that CD8+ T cells are an important target for cancer therapy ^13-15^. Resting memory CD4 T cells^16 17^ and M0 macrophages are usually recognized as nonactive cells^18^. CD8+ T cells were middle collelation with the two cells; this indicated the activation of the two cells were associated with CD8+ T cells.

**Figure 2.**
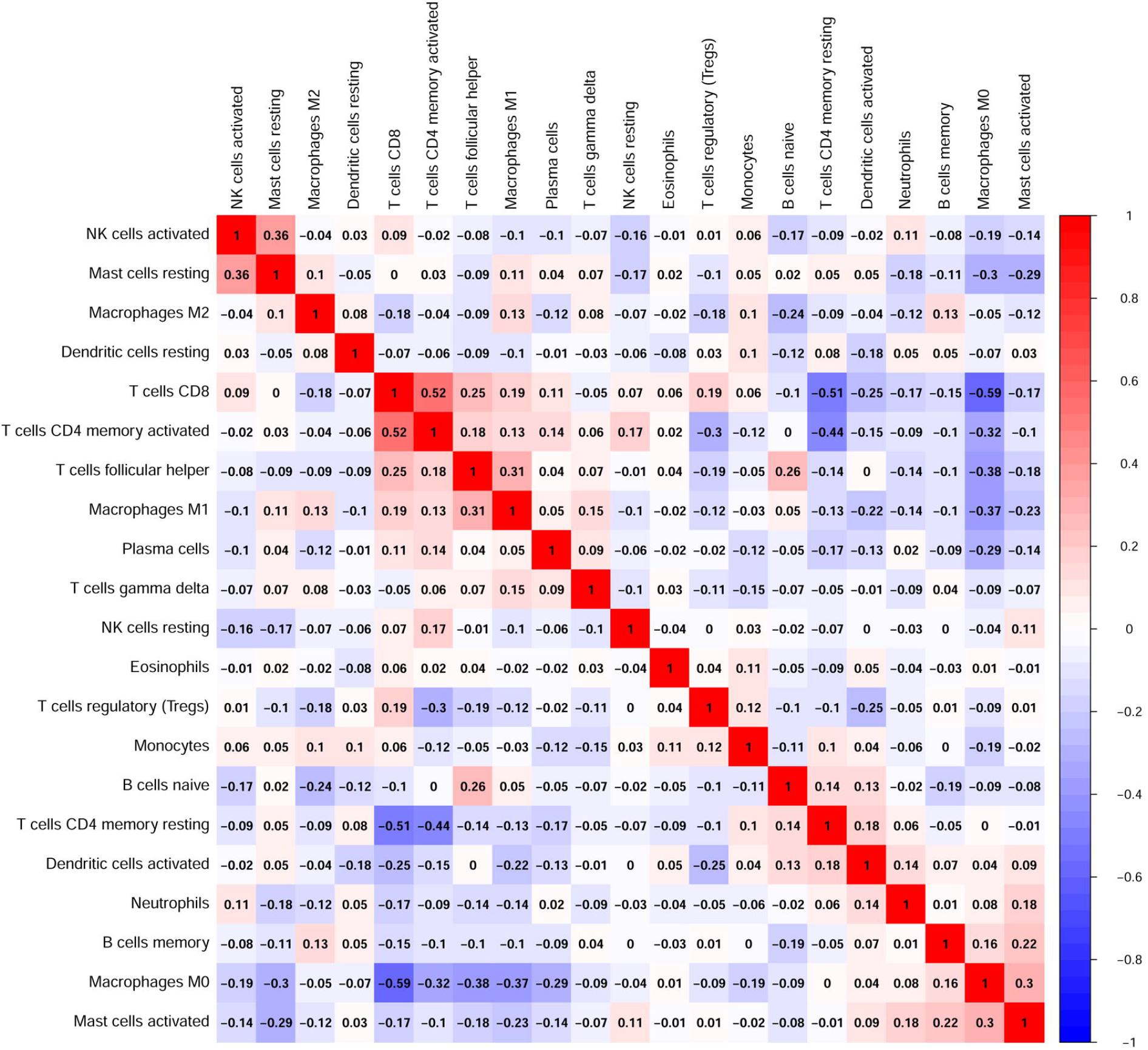
The correlation of cytolytic activity in TCGA.

### Association of TIICs with EMC cell differentiation grade and EMC invasion and metastasis

An interesting result was that the proportion of resting dendritic cells, resting memory CD4 T cells, and Tregs decreased according to the cancer cell differentiation (G); in contrast, the proportion of memory-activated CD4 T cells and M1 and M2 macrophages increased according to the cancer cell differentiation grade (Figure 3). Resting dendritic cells and resting memory CD4 T cells are usually considered as nonactive immune cells^19 20^, while Tregs were found to inhibit endogenous immune responses against tumors^21^. These immune cells might be involved in EMC carcinogenesis; these cells involved in other cancer development was also reported previously^22^. Memory-activated T cells and M1 and M2 macrophages are usually recognized as active immune cells, and our results showed that these cells also play a role in tumor cell differentiation.

**Figure 3.**
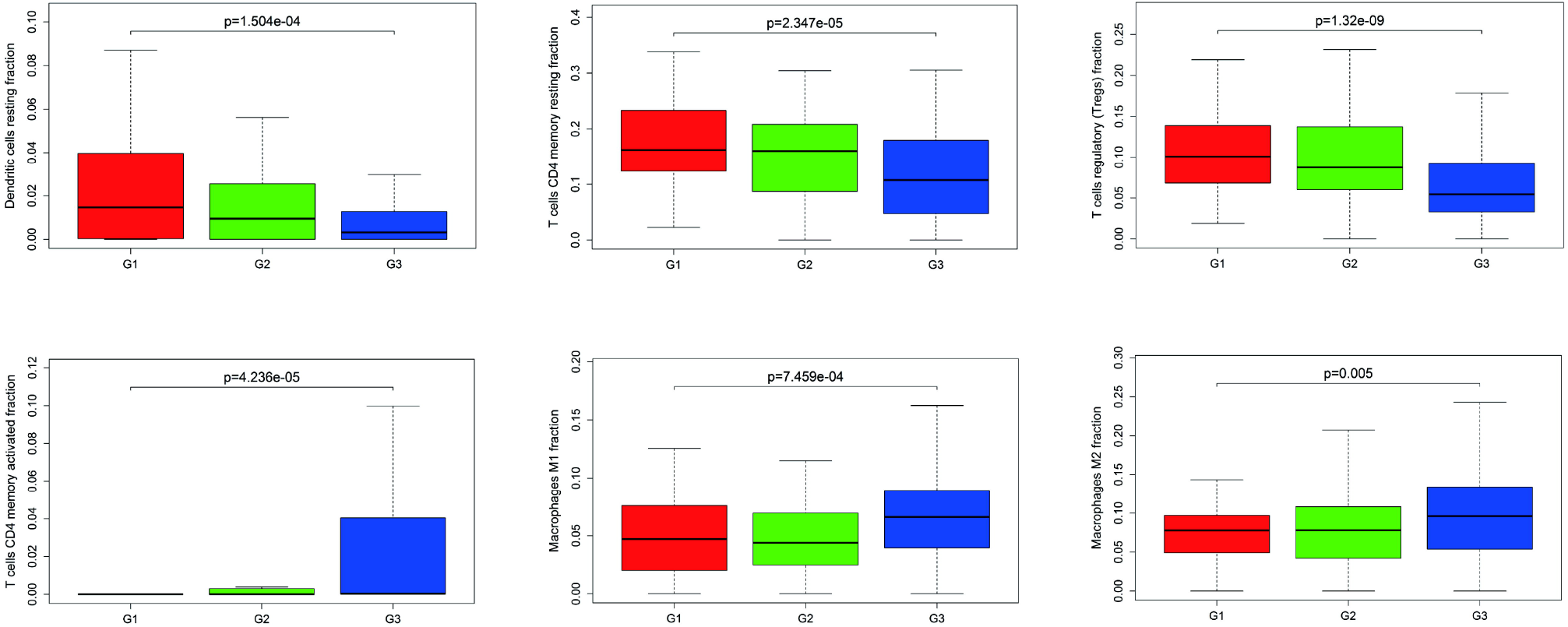
The proportion of TIICs according to the cell differentiation grade of EMC.

We also analyzed the correction between TIICs and regional lymph node (N) involved, and did not find significant TIICs differences in EMC N0, N1, and N2 (Table S1).

### Identification of prognostic subsets of TIICs in different clinical stages of EMC

We assessed whether there was a potential correlation between TIICs and patients’ clinical stage (FIGO). It was found that the proportion of Tregs decreased during EMC development (Figure 4A), while the proportion of gamma delta T cells increased in advanced EMC (Figure 4B). A diverse monocyte population was observed according to the clinical stages (Figure 4C). Tregs, a subpopulation of suppressive T cells, are potent mediators of self-tolerance and are essential for the suppression of triggered immune responses^23^; a decrease in Tregs during EMC development indicates an increase in anticancer activity. T cells gamma delta are a positive immune activity indication for cancer therapy^24 25^, and also a positive factor in EMC. Monocytes and monocyte-derived macrophages play key roles in tumor progression^26^. Hanna *et al* reported that patrolling monocytes blocked tumor access to the lung^27^. A diverse range of monocytes in the different EMC stages indicated their multifunctionality in EMC metastasis, but further investigations are required to confirm this speculation.

**Figure 4.**
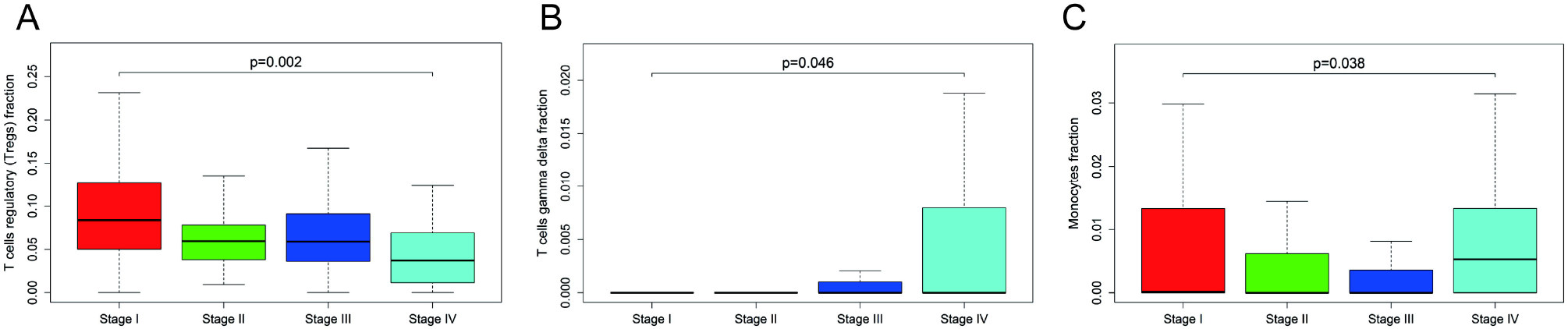
The proportion of TIICs according to the clinical stages of EMC.

### Subsets of TIICs in pre- and postmenopause patients with EMC

Because female sex hormones are present in different levels in pre- (Pre) and postmenopause (Post) women^28-30^, we assessed whether TIICs were affected by hormones. We found that the proportion of naïve B cells (Figure 5A) and resting memory CD4 T cells (Figure 5B) decreased in the Post group. Because both these cell populations are recognized as nonactive immune cells in cancer^20 31^, the decrease in their proportion might be influenced by hormone levels; this aspect requires further investigation.

**Figure 5.**
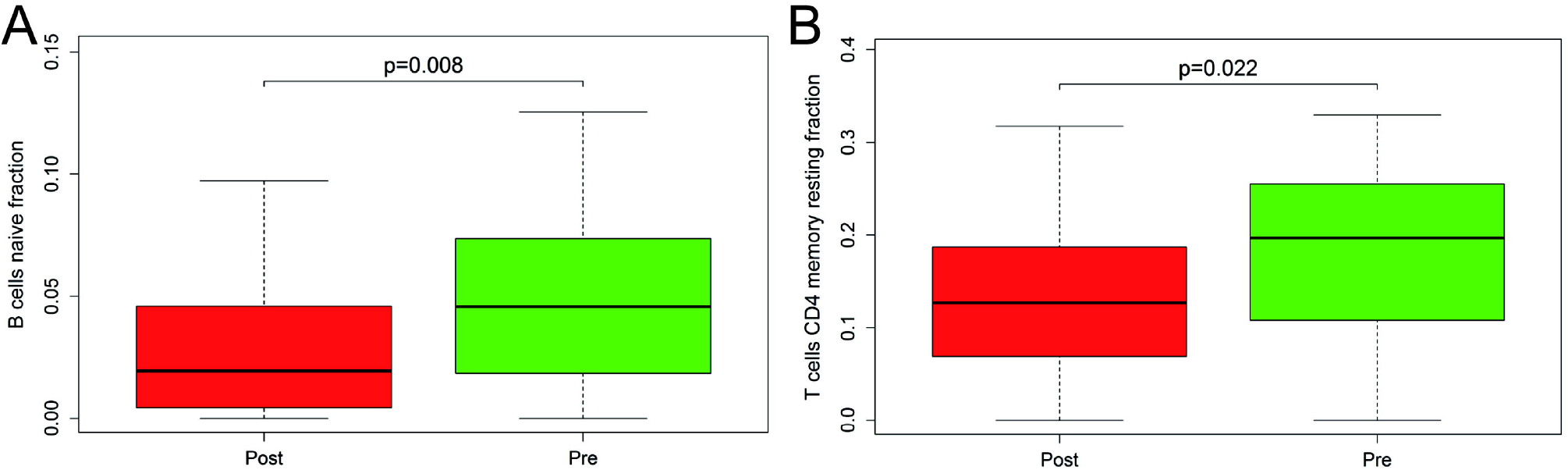
Subsets of TIICs in pre (Pre) and postmenopause (Post) patients with EMC.

### Identification of prognostic subsets of TIICs in EMC patients’ survival time

We found that a lower proportion of activated dendritic cells (Figure 6A) and gamma delta T cells (Figure 6B), and higher T regulatory cells (Tregs) (Figure 6C) predicted a longer EMC survival time than their opposites. Dendritic cells have the potential to overcome tumor tolerance and induce antitumor immunity when loaded with tumor antigens^32^. Despite high potential in promoting antitumor responses, tumor-associated DCs are largely defective in their functional activity and can contribute to immune suppression in cancer^32^. Our present results showed that a high proportion of activated dendritic cells was associated with poor prognoses of patients with EMC. This finding implied that an effective approach to treat EMC was to decrease the activation of dendritic cells. Because gamma delta T cells have potent cytotoxicity and can produce interferon-γ, they are considered to play a protective role in cancer^33^. Furthermore, these cells were reported to be poor prognostic biomarkers in human breast cancer^34^, and we found a similar property of these cells in EMC. Interestingly, Tregs have been shown to exhibit antitumor properties in the tumor microenvironment^35^. We found that a higher proportion of Tregs could improve EMC prognosis, especially in the first 10 years of follow-up.

**Figure 6.**
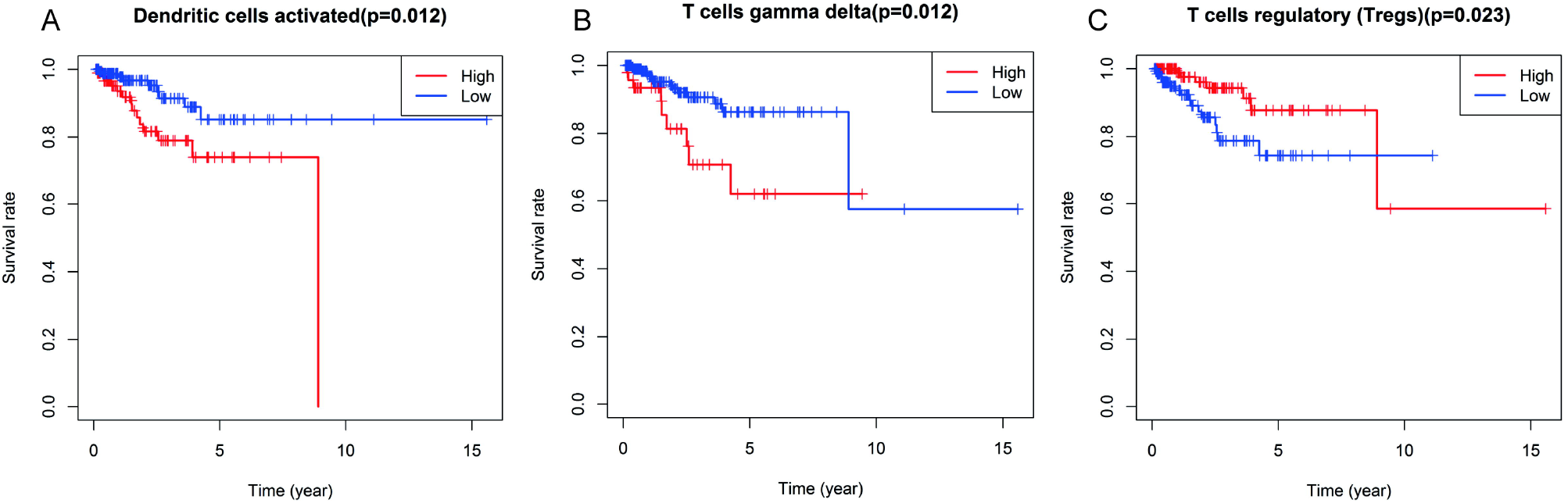
Effect of subsets of TIICs on the survival time of patients with EMC.

### Identification of subsets of TIICs in therapy-responding EMC patients

Based on the TCGA patients’ clinical data, we found that patients with low gamma delta T cells had more effective responses than the patients with high gamma delta T cells (Figure 7).

**Figure 7.**
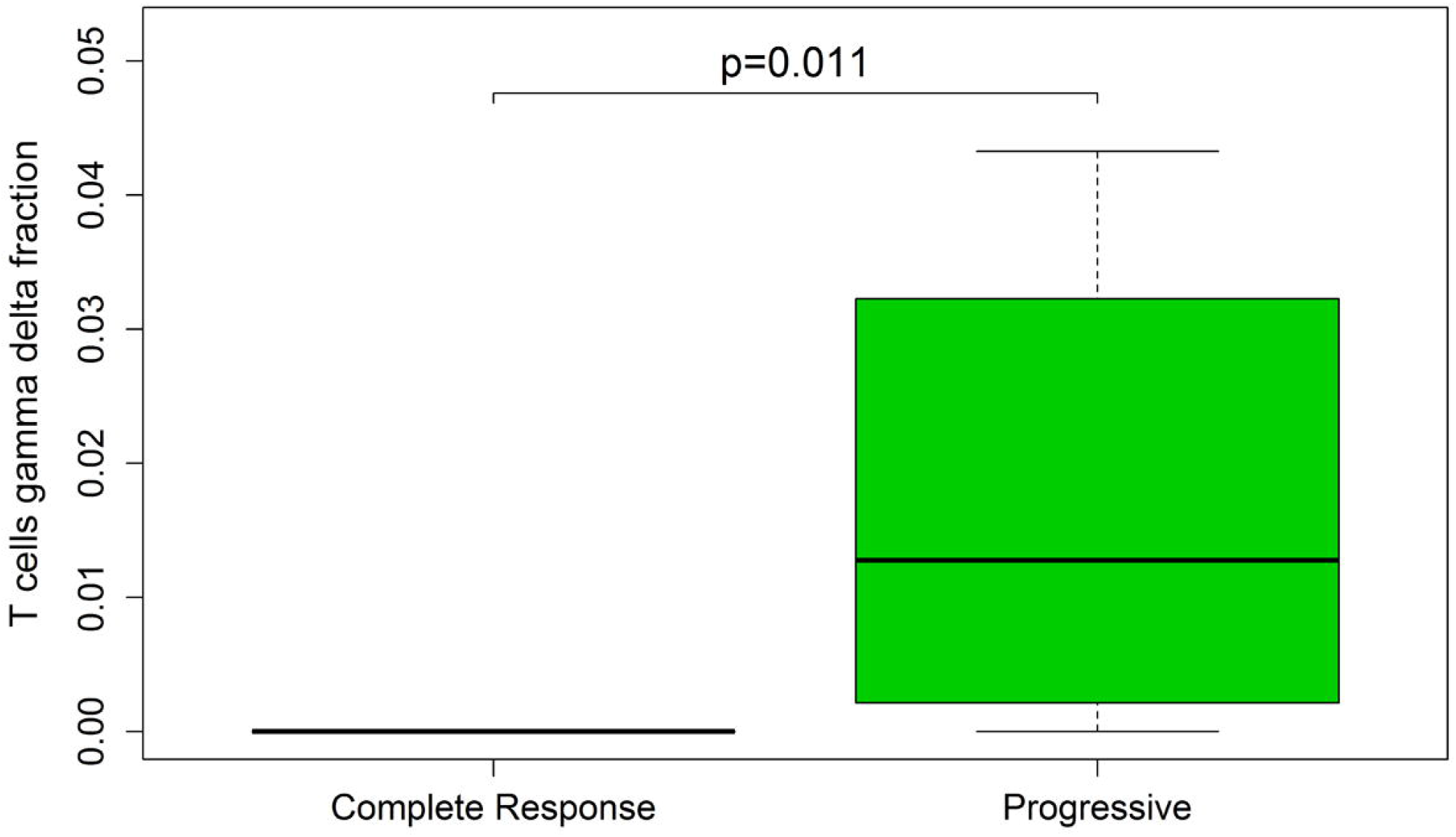
Gamma delta T cells were associated with response to therapy in patients with EMC.

## Discussion

Recently, CIBERSORT, a computational method for analyzing cell populations, has been used to define the proportion of 22 types of TIICs in tissues^36^. This method is based on the genome transcripts of the 22 types of TIICs, and the results of TIICs were combined with patients’ clinical data to systematically analyze the association between the proportion of TIICs and patient characteristics^36^. Thus, this approach could overcome the limitation of traditional immunohistochemistry (IH)-based methods because IH defines cell type mainly on the basis of one or two cell markers.

EMC is a hormone-related cancer^37^. The most common lesions (type 1) are typically hormone sensitive and low stage, and they have an excellent prognosis^37^. Hormones can significantly affect the immune system^38^. Our present study revealed that the proportion of naïve B cells and resting memory CD4 T cells were different in pre- and postmenopause women. Thus, the association between TIICs and hormone levels needs further investigation.

Interestingly, the EMC patients with high gamma delta T cells shown low responders to therapy, and with low gamma delta T cells had longer survival times. This result indicated gamma delta T cells might be a key factor for EMC treatment.

The immunotherapy of cancer has made some significant advances in the past few years, with some favorable results^39^. A recent study showed that a combination of immunomodulation, CARs, and immunotherapy might be the next direction for cancer immunotherapy^40^. In the present study, we indicated potential immune cells as targets for the therapy of EMC on the basis of TCGA datasets. CIBERSORT was used only to estimate the proportions of TIICs. Each TIIC has multi-subsets, and each of the TIIC subsets has wide functions. The position of TIICs and their interaction with different microenvironments would have different effects^41^. Thus, more studies are needed to completely under

## Data Availability

All related data are available if needed.

## ACKNOWLEDGMENTS

We thank Dr. Jianfeng Li for the help in Statistical analyse

